# Global access to speech hearing tests

**DOI:** 10.1101/2024.06.12.24308842

**Authors:** Sigrid Polspoel, David R. Moore, De Wet Swanepoel, Sophia E. Kramer, Cas Smits

## Abstract

Understanding speech in noisy settings is one of the biggest challenges for individuals with hearing loss. Traditional speech-in-noise tests play a crucial role in screening for and diagnosing hearing loss, but are resource-intensive to develop, limiting accessibility, particularly in low and middle-income countries. This four-part study introduces an innovative approach using artificial intelligence (AI) to automate the development of such tests. By leveraging text-to-speech (TTS) and automatic speech recognition (ASR) technologies, this approach significantly reduces the cost, time, and resources required for high-quality speech-in-noise testing accessible worldwide. The procedure, named “Aladdin” (Automatic LAnguage-independent Development of the Digits-In-Noise test), creates digits-in-noise (DIN) hearing tests through synthetic speech material and ASR-based level corrections to perceptually equalize the digits, demonstrating characteristics comparable to traditional tests. Notably, Aladdin provides a universal guideline for developing DIN tests across languages, addressing the challenge of comparing test results across variants. This approach, with its potential for broad application in audiology, represents a significant advancement in test development and offers a cost-effective and efficient enhancement to global screening and treatment for hearing loss.

## INTRODUCTION

The ability to understand speech in noisy environments is the most common challenge for individuals with hearing loss, impacting daily communication and quality of life. Traditional speech-in-noise tests, essential for assessing the ability to recognize speech in noise, diagnosing hearing loss and evaluating hearing aid and cochlear implant performance, require extensive resources for development. This limits their availability, especially in low and middle-income countries. The present study introduces an innovative approach using artificial intelligence (AI) to automate the development of such tests. By employing text-to-speech and automatic speech recognition technologies, this approach drastically reduces the cost, time and resources required to develop high-quality speech-in-noise testing accessible worldwide.

Since the establishment of the field of audiology, the importance of using speech-in-noise tests has been recognized ^1^. Speech-in-noise tests measure an individual’s ability to recognize speech by presenting speech stimuli (usually lists of digits, words or sentences) in the presence of noise. The outcome of the test is either a percentage-correct score or the signal-to-noise ratio (SNR) where the average speech recognition score is equal to a certain percentage-correct ^2^. Typically, the listener is presented with a series of speech stimuli and is asked to repeat what they heard. One example of such a test, and the main focus of this study, is the widely used digits-in-noise (DIN) test ^3,4^. It uses digit triplets presented in speech-shaped noise to measure the speech recognition threshold (SRT). The SRT is the signal-to-noise ratio, expressed in dB SNR, at which the listener recognizes 50% of the digit triplets correctly. Apart from a diagnostic test in clinics ^5,6^, the DIN is used as a quick self-test for hearing screening that can be completed via internet or a smartphone app ^6–9^, or as a home test for cochlear implant users ^10^. The test results highly correlate with standard audiometric findings ^11^ and can reach a large global audience without the need of an examiner ^11–13^ or the requirement to calibrate the devices at home ^14,15^. The World Health Organization has adopted this test for its official hearing screening app, called hearWHO ^16^, and included the DIN test as a recommended option for hearing screening in adults and school-aged children ^17^.

An essential property of a speech-in-noise test is the precision of the test and, for use as a screening test, its specificity and sensitivity. The precision of the SRT estimated in a speech-in-noise test is expressed by the standard error of measurement (SEM). A small SEM is necessary to produce valid and reliable speech-in-noise test results. The SEM is inversely proportional to the slope of the speech recognition function of the test ^18^, a sigmoidal function representing the percentage correctly repeated speech items against the SNR at which they are presented. The speech recognition function is described by the SRT (i.e., SNR at 50% correct; the midpoint of the function), and the slope of the speech recognition function at the midpoint. Because each test involves a series of different stimuli, the average speech recognition function is formed by the speech recognition functions of the individual stimuli ^19^. Differences in intelligibility between the stimuli make the slope of the average speech recognition curve shallower and therefore increase the SEM. Thus, ensuring perceptual equivalence among individual speech stimuli is important as it steepens the speech recognition function and minimizes SEM. For hearing screening tests, additional properties determine the quality of a test: sensitivity refers to the ability of the test to identify those with hearing loss and specificity refers to the ability of a test to identify those without hearing loss.

Making the speech stimuli perceptually equivalent is a crucial aspect in the development of an accurate and precise speech-in-noise test. This is generally a costly and labor-intensive process (Figure 1 [A]). It involves the professional recording of speech stimuli uttered by trained speakers and selecting the best utterances. Then a listening experiment is conducted with listeners with normal hearing to use the results for selecting or creating perceptually equivalent stimuli ^20,21^. The need for trained speakers, listeners with normal hearing, professional recording equipment, audiometric equipment, and a sound attenuating booth significantly limits the development of new speech-in-noise tests, especially in low and middle-income countries. The aim of the current study was therefore to develop a procedure for the automatic development of speech-in-noise hearing tests using AI (Figure 1 [B]).

**Fig. 1.**
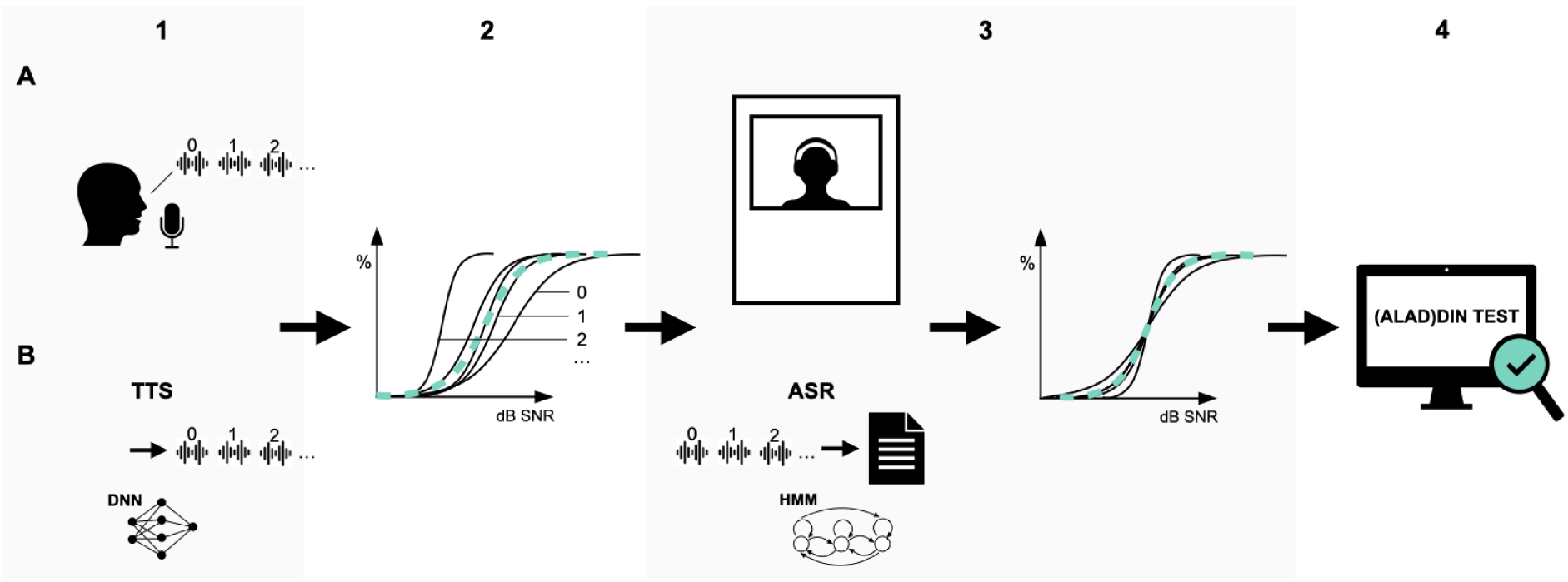
Schematic representation of the traditional development process of the DIN test’s speech material (A) versus the automated process Aladdin (B). The columns numbered 1 to 4 correspond to Part 1 to 4 of this article. **A: (1)** Digits are recorded by a professional speaker and the best recordings are selected. **(2)** Speech recognition functions of the recorded digits differ in slope and SRT, i.e., the intelligibility of the digits is not equal. The blue dotted line represents the average speech recognition function of the digits. **(3)** A listening experiment with listeners with normal hearing is set up to determine the level corrections per digit needed to achieve equal intelligibility. Next, **l**evel corrections are applied, horizontally shifting the speech recognition functions to the mean SRT. Now all digits have the same expected SRT, and the average speech recognition function (blue) has become steeper than in step 2, resulting in a smaller SEM. **(4)** The test is evaluated with groups of listeners with normal hearing and hearing loss**. B:** The automated process, “Aladdin”, replaces all the steps of the traditional development process. **(1)** A deep neural network (DNN)-based text-to-speech (TTS) system creates digit recordings. **(3)** Level corrections are determined using the output of a Hidden Markov model (HMM)-based automatic speech recognition system (ASR). The corrections are subsequently applied to the individual digit recordings.

In the current study, the procedure of creating a DIN test was drastically shortened and simplified by AI techniques. Specifically, text-to-speech (TTS) and automatic speech recognition systems (ASR) were deployed to create DIN tests. TTS was used to create the synthetic speech material (digits) and ASR to determine level corrections needed to achieve perceptual equality among the synthetic digits (Figure 1 [B]). We refer to this as ‘Aladdin’; “Automatic LAnguage-independent Development of the Digits-In-Noise test”. Aladdin’s goal is to provide a simple, efficient, automatic, low-cost and universal way to create new DIN tests in different languages.

Aladdin requires a high-quality TTS system which, thanks to recent advances in deep neural network (DNN) techniques, can now approach the naturalness of human speech ^22^. Synthetic speech has emerged as a successful alternative to natural speech for creating speech material in various languages, as demonstrated in studies on speech-in-quiet ^23^ and speech-in-noise tests ^24–28^. In addition to a TTS system, Aladdin requires ASR to determine level corrections per digit needed to equalize intelligibility among the digits. Two types of ASRs were considered: off-the-shelve, pre-trained systems with large language models; and a specific system that was trained on the speech stimuli of choice.

There were four parts to this Aladdin study. **Part I** – Creating synthetic speech material in five languages and assessing its subjective speech quality. The speech quality of synthetic material and original speech material was measured using the 5-point mean opinion score (MOS) scale. Assessment was done by native listeners in their own country via a calibrated laptop. The goal was to determine if the subjective quality of synthetic speech material was comparable to reference speech material used in audiometric testing. If so, we can conclude that synthetic speech is a viable option for creating speech material for speech-in-noise tests. **Part II** – Comparing speech recognition functions of natural and synthetic digits. A listening experiment was conducted to determine speech recognition functions for digit triplets using male and female Dutch synthetic voices, and a male Dutch natural voice (reference Dutch speech material). SRTs, slopes and variability of the natural and synthetic digit recognition functions were compared. Results yielded level corrections per digit (0 - 9), useable as a human reference for the ASR. **Part III** - Feeding natural and synthetic digits in noise to different ASRs to determine level corrections for each digit automatically. These level corrections were compared to those based on human listeners from Part II. A strong correlation between the level corrections, as well as a low mean squared error between the human and TTS based level corrections, would suggest that the human listening experiment can be replaced by an ASR. The ASR that resembled the human performance the most was used for the remainder of the study. Next, we applied the ASR-based level corrections to the synthetic digits, making the digits perceptually equally intelligible. At this point, the speech material of the Aladdin-test was created. **Part IV** – Evaluation of the Aladdin test in listeners with normal hearing and listeners with hearing loss. Both groups performed the Aladdin test in Dutch and English, as well as a reference DIN test in both languages. The outcomes and screening characteristics were compared to evaluate whether the Aladdin tests are valid alternatives to the reference DIN tests that were developed the traditional way.

## RESULTS

### Part I: Subjective quality of synthetic (text-to-speech) speech material for hearing tests

Generating speech material is the first step of developing a speech-in-noise test (Figure 1 [B1]). Google Cloud TTS ^29^ was used to generate synthetic speech in a male and female voice in 5 languages: Dutch, English, French, Spanish and Mandarin. The speech material included the same digits, words and sentences that are used in standard speech audiometry in countries for which each language is the most common. Rating took place locally in the countries by 10 native-speaking participants per language. They were provided with a calibrated laptop with self-explanatory software to complete the speech rating task. Note that this experiment was solely utilized for the evaluation of this phase of the Aladdin test development procedure. It will not be required to conduct this phase in future developments of Aladdin tests in different languages. The sound files were utterances of digits, words and sentences presented in quiet and in low-levels of speech-shaped masking noise in each of 4 voices: a natural voice (male or female), a synthetic male voice, a synthetic female voice and a degraded voice (i.e., vocoded sound file). The question they were asked after listening to each sound file was: “Overall impression - how would you rate the quality of the speech you just heard?”. In cases where the speech was presented in background noise, “(ignore the noise)” was added to the sentence. Participants would then rate the speech quality based on a MOS scale^30^, which is composed of 5 labelled points from 1-Very poor to 5-Excellent. Each participant was presented with each type of voice 3 times. MOS scores averaged across participants and repetitive presentations are shown in Figure 2.

**Fig. 2.**
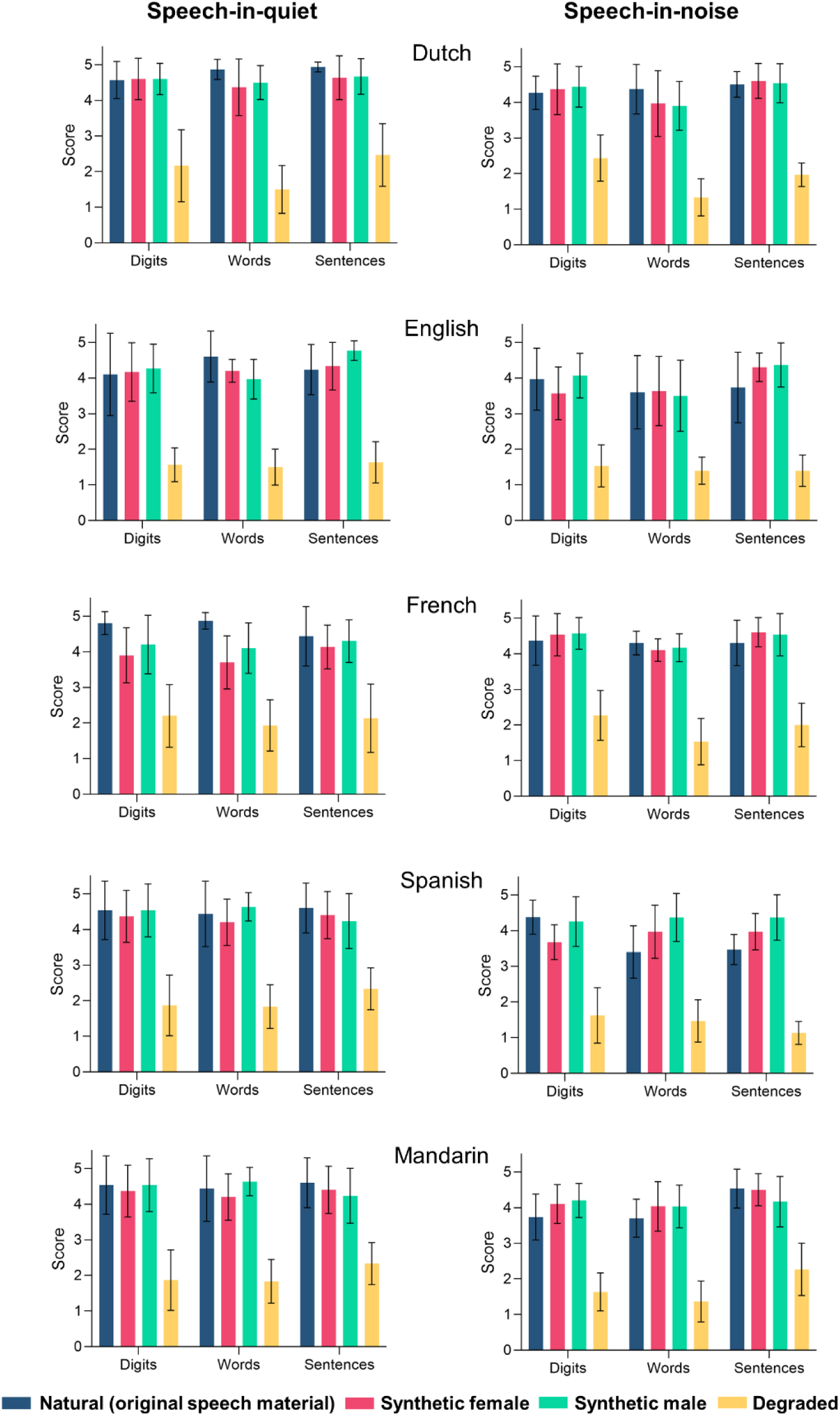
Average MOS scores for three types of speech material (digits, words and sentences) for a natural, synthetic female, synthetic male and degraded voice in quiet (left) and in noise (right). Scores are the means (± SD.) of 10 participants per language.

The majority (16 out of 18) of the synthetic and natural speech sound files received an average MOS score above 4, meaning that all voices, except for the degraded one, were considered to be of good to excellent speech quality. Across languages, there were no significant differences between the male and female synthetic voices and the natural voice in either the digits (χ²(2)=0.05, *p* = 0.98), words (χ²(2)=0.68, *p* = 0.71), or sentences (χ²(2)=0.54 *p* = 0.77) material in quiet. Similarly, there were no significant differences observed in noisy conditions (digits: χ²(2)=1.23, *p* = 0.54; words: χ²(2)=1.27, *p* = 0.53; sentences: χ²(2)=0.22, *p* = 0.89). The degraded voice was, as expected, consistently rated the worst. The results suggest that synthetic speech material is not only suitable for speech-in-noise tests, but also for speech-in-quiet tests. A hypothesis was that the synthetic voice would be rated worse on the sentence material compared to the digits material, because longer speech fragments with more prosody and co-articulation could reveal subtle unnatural sounds in the synthetic voice. However, since ratings of synthetic and natural voices did not differ in any of the three speech materials, it appears that synthetic speech can be effectively used for both digits-in-noise tests and tests involving other words or sentences.

### Part II: Comparing speech recognition functions of natural and synthetic digits

Further potential differences between the natural and synthetic voices were explored by comparing speech recognition functions of individual digits (Figure 1 [2]). To do so, speech recognition functions of the synthetic male, the synthetic female and the natural male Dutch digits from Part I were determined by presenting the digits at fixed SNRs to a group of listeners with normal hearing (n = 24). The mean (± standard deviation) SRTs and mean slopes of the digits were -9.4 ± 3.2 dB SNR and 14.9 ± 4.1 %-points/dB respectively for the natural voice, -8.6 ± 3.3 dB SNR and 18.1 ± 6.0 %-points/dB for the female synthetic voice, and -8.9 ± 3.7 dB SNR and 16.6 ± 9.9 %-points/dB for the male synthetic voice (Figure 3). These mean SRTs were not significantly different (ANOVA, *F*(2,27) = 0.144, *p* = 0.86), and neither were the mean slopes (ANOVA, *F*(2,27) = 0.51, *p* = 0.61) suggesting that digit intelligibility and change in intelligibility with SNR for synthetic digits are within the variability range of those for natural digits.

**Fig. 3.**
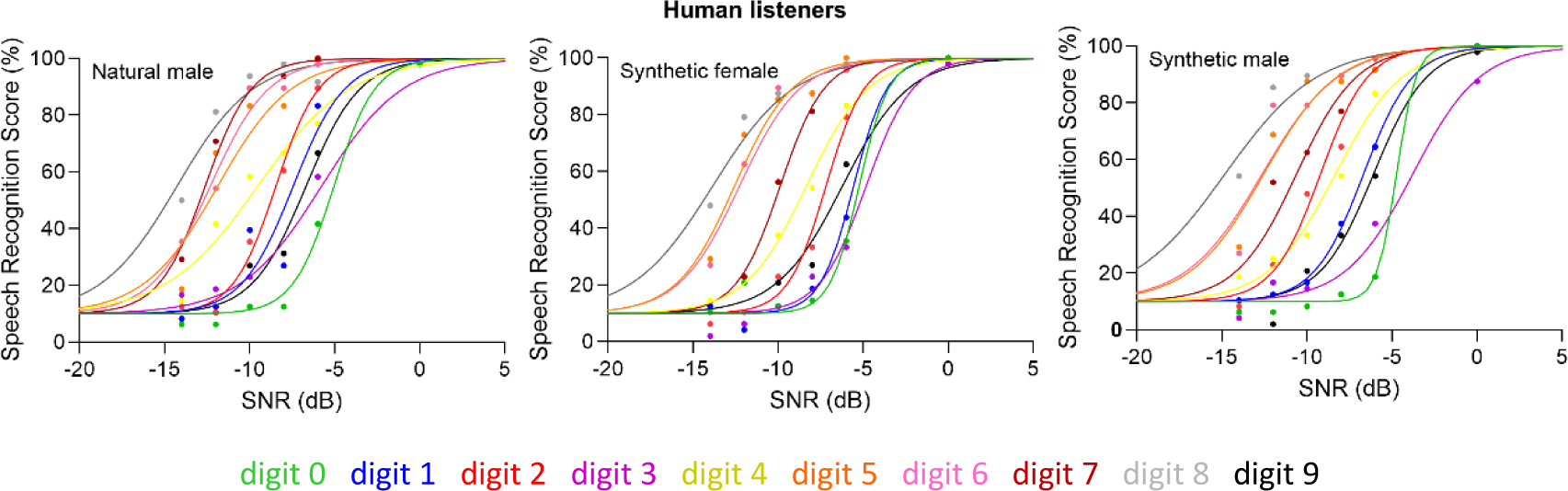
Speech recognition functions (percent correct as a function of SNR) of the individual digits in noise for normal hearing listeners where the digits are uttered in 3 voices: a natural male (left panel), a synthetic female (middle panel) and a synthetic male (right panel) voice. Each dot shows the average score of 24 participants. The lines show the maximum likelihood fits to the raw data.

The level corrections per digit needed to achieve perceptual equivalence were calculated by subtracting the (voice-specific) mean SRT from the digit-specific SRT. Level corrections shifted the individual speech recognition functions horizontally so that all SRTs aligned on the average SRT (see Figure 1). The level corrections of the natural male and synthetic male voice were strongly correlated (Pearson, r = 0.96, *p* < 0.001) (Figure 4). There was also a strong correlation between level corrections of the natural male and synthetic female voice (Pearson, r = 0.95, *p* < 0.001). Strong correlations among the voices imply that the level corrections are robust to type of voice. These level corrections, obtained from human listeners, serve as a benchmark for the ASR in Part III.

**Fig. 4.**
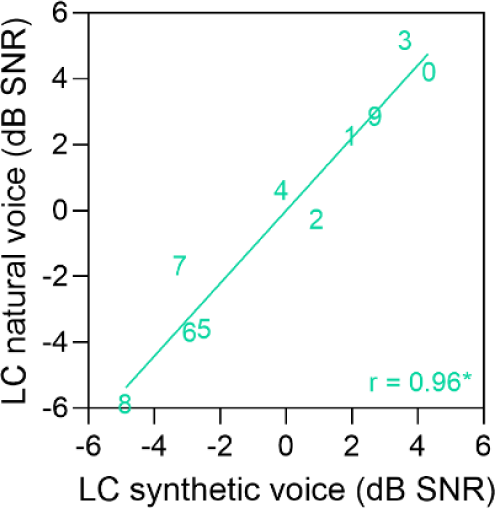
Level corrections (LC) for the natural male digits as a function of the LC of the synthetic male digits based on a listening experiment with 24 human listeners.

### Part III: Determining level corrections from ASR

Part III focused on generating speech recognition functions per digit with various ASRs to determine level corrections needed to perceptually equalize intelligibility (Figure 1 [3B]). If both speech recognition functions and the resultant level corrections from an ASR closely resemble those produced by human listeners, then the ASR could serve as a satisfactory alternative for conducting listening experiments in development of DIN tests. First, three off-the-shelf cloud-based ASRs were considered: Google Cloud ASR ^31^, Microsoft Azure Bing Speech API ^32^ and IBM Watson Speech to Text ^33^. Dutch digit triplets at various SNRs ranging from -4.5 to 7 dB in steps of 0.5 dB were presented to the three systems and the written output was cleaned and scored. The reason for using such small SNR increments was motivated by the discovery of a distinct tipping point, where the digits were either recognized by the ASR or not. This resulted in very steep speech recognition functions (Figure 5 [B-D]). For comparison, speech recognition functions of the same synthetic female digits derived from human listeners (see Part II) are shown in Figure 5 [A]. The average human-based SRT was much lower than the average ASR-based SRTs (one-way ANOVA, *F*(3,36) = 39.934, *p* < 0.001), and the average slope was much more gradual in the former (one-way ANOVA, *F*(3,36) = 4.002, *p* = 0.015). The variance of the digit SRTs was significantly greater for humans compared to the ASRs (Levene’s test, *F*(3,36) = 7.465, *p* < 0.001). This indicated that the digits were more equally intelligible for ASRs than for humans. The notably higher SRTs and steeper slopes in ASR-generated speech recognition, along with moderate correlations (Pearson, 0.54 ≤ r ≤ 0.79) between ASR-derived and human-evaluated level corrections (see supplementary material), suggested that these pre-trained ASRs were not suitable for Aladdin’s intended purpose.

**Fig. 5.**
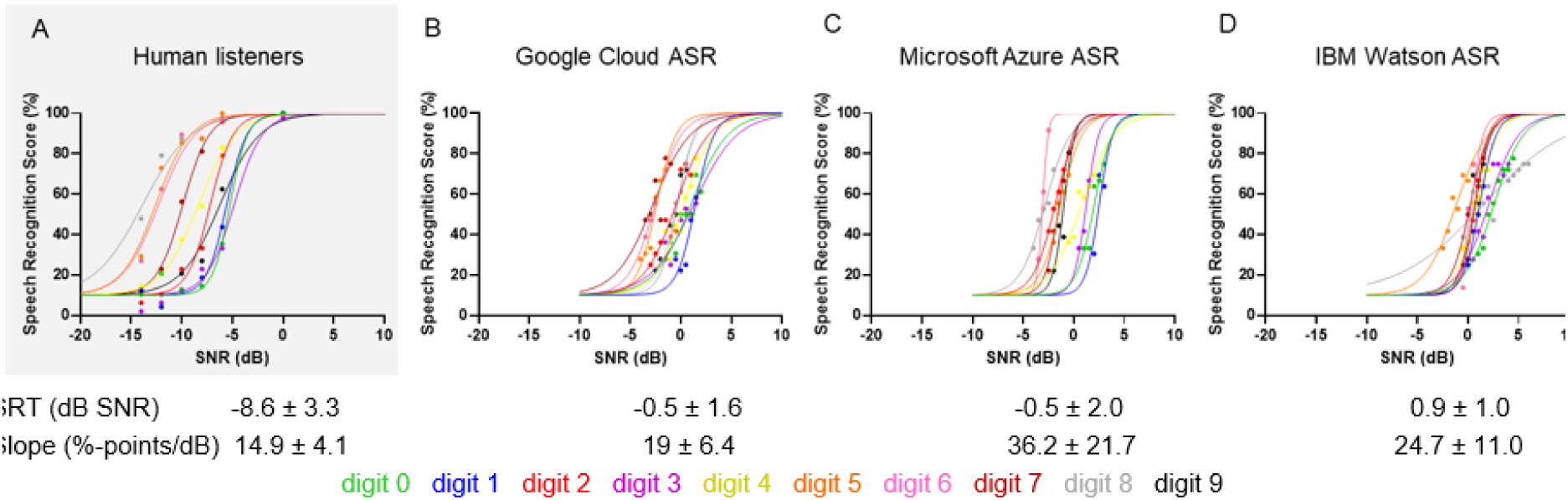
Speech recognition functions of the synthetic female Dutch digits in noise based on three cloud-based ASRs: Google Cloud ASR (B), Microsoft Azure Bing Speech API (C) and IBM Watson Speech to Text (D). A: Reference human-derived speech recognition functions of the same digits. The dots are the percentage correct scores based on 12 presentations per digit per SNR. The lines show the maximum likelihood fits to the raw data.

A specific ASR called FADE, developed by Schädler, et al. ^34^, was designed to forecast human auditory performance in speech recognition tasks. This ASR is based on Gaussian mixture models and hidden Markov models (GMM-HMM) that simulates the human speech recognition process ^35,36^. Unlike the off-the-shelf (standard) ASRs mentioned earlier, this particular system employs identical speech and noise stimuli for both training and testing purposes. As a result, it yields much lower SRTs that closely align with those of humans ^35^.

FADE was trained on the noisy digits speech material at various SNRs, separately for natural Dutch, synthetic Dutch and synthetic English digits. Subsequently, speech recognition functions were generated by testing speech recognition of the digits at various SNRs by the trained models. Note that FADE works independently of languages and the Aladdin approach is also language independent. After a first run through FADE (Part 1), level corrections were determined and applied to the digit sound files. The level corrected digit sound files were again run through FADE (Part 2) and new level corrections were determined and applied on top of the previous corrections. This process was repeated until the slope of the average speech recognition function did not increase more than 1%-point/dB. There was a total of five runs for the Dutch natural digits, three runs for the Dutch synthetic digits, and two runs for the English synthetic digits. Figure 6 shows the average speech recognition functions after the different runs. It is evident that, as anticipated, the application of level corrections resulted in steeper speech recognition functions.

**Fig. 6.**
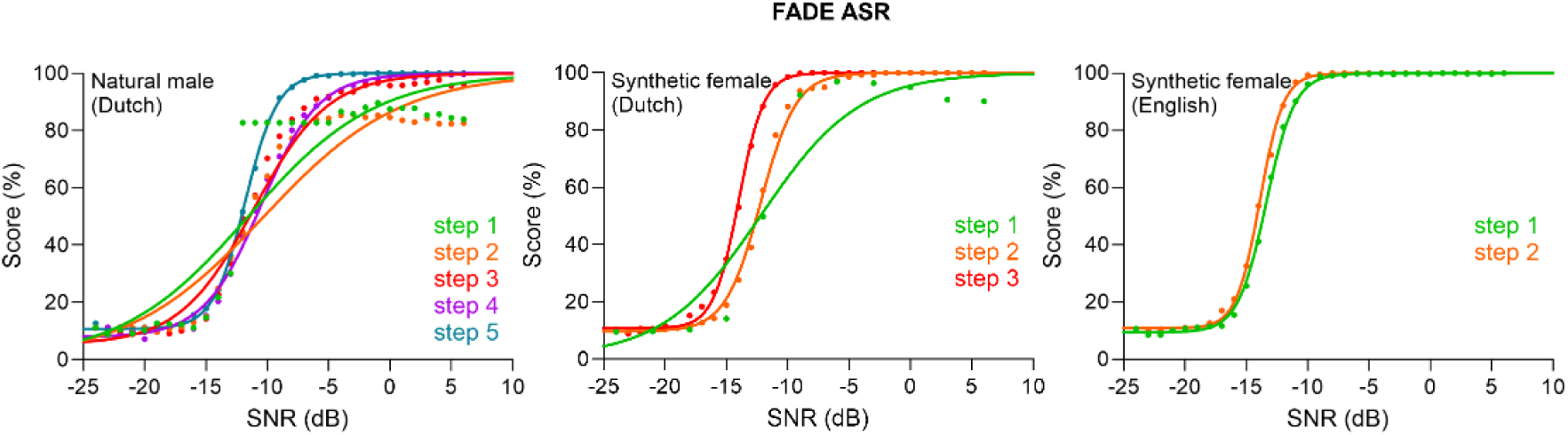
Average speech recognition functions of the natural male Dutch digits, the synthetic female Dutch digits and the synthetic female English digits based on the ASR ‘FADE’. The different functions are the result of the different runs through the system. After each run, level corrections were determined to achieve equal intelligibility and these level-corrected digits were again fed to the system until the slope stopped increasing more than 1%-point/dB.

The cumulative total of level corrections across various runs for each digit, meaning the corrections applied to the final set of digits, was used to compare with the reference human-based level corrections determined in Part II. Note that we had a reference only for Dutch natural and synthetic digits (male voice), not for the English synthetic ones. Figure 7 shows strong correlations between human and ASR-based level corrections, for both natural (Pearson correlation, r = 0.88, p < 0.001) and synthetic (r = 0.96, p < 0.001) digits. Root mean square (RMS) error of the level corrections was 1.6 dB for the natural voice and 0.9 dB for the synthetic voice.

**Fig. 7.**
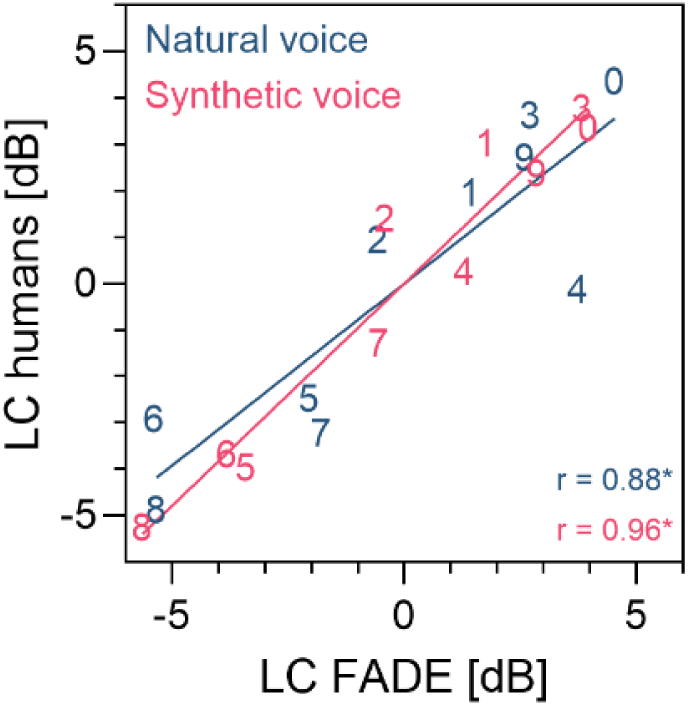
Level corrections (LC) for natural and synthetic Dutch digits based on the listening experiment with human listeners from Part I as a function of the LC based on the FADE ASR.

### Part IV: Comparing Aladdin to original DIN test

The final set of level-corrected Dutch and English synthetic digits from Part III were used to create new Dutch and English DIN tests, referred to as ‘Aladdin tests’. In Part IV, Aladdin tests were validated (Figure 1 [4]) and compared to established reference Dutch and English DIN tests. Two groups participated in the study: native Dutch-speaking listeners with normal hearing (n = 28) and those with hearing loss (n = 20, with a mean pure tone average threshold (PTA) at 0.5, 1, 2, and 4 kHz in the better ear of 47 ± 19 dB HL). Each group performed 4 DIN tests: the reference Dutch DIN test, the Dutch Aladdin test, the reference English DIN test and the English Aladdin test. They did each test twice (test and retest). All participants had at least a self-reported basic understanding of the English language. Figure 8 shows the speech recognition thresholds (SRTs) of the Aladdin test as a function of the SRTs of the reference DIN test in Dutch (left panel) and English (right panel). The circles and squares represent the test-retest SRT averages of the participants. There is a strong correlation between the Aladdin SRTs and the reference DIN SRTs for both the Dutch (Pearson correlation, r = 0.91, p < 0.0001) and English (r = 0.95, p < 0.0001) versions across all listeners.

**Fig. 8.**
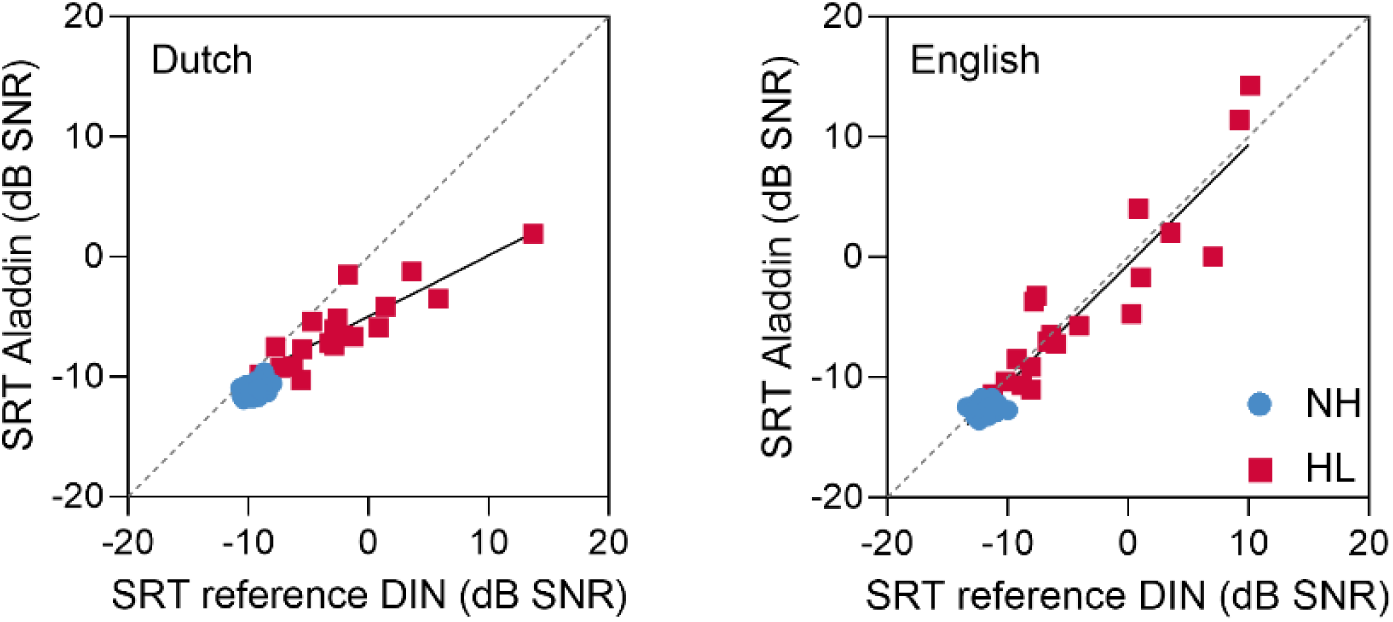
Speech recognition thresholds (SRTs) of the Dutch (left panel) and English (right panel) Aladdin test as a function of the reference DIN test. The blue circles represent the listeners with normal hearing (NH), the red squares the listeners with hearing loss (HL).

Figure 9 illustrates the test-retest correlation of SRT scores for the four DIN tests. The Standard Error of Measurement (SEM) for the listeners with normal hearing and the listeners with hearing loss combined was 1.4 dB for the Dutch reference DIN, 0.6 dB for the Dutch Aladdin, 1.5 dB for the English reference DIN, and 1.7 dB for the English Aladdin. In the group of listeners with normal hearing, the SEM was 1.0 dB, 0.4 dB, 0.5 dB, and 0.5 dB for the four tests, respectively. The SEM represents the measurement error of the SRT estimate. As anticipated, the SEMs were higher in the listeners with hearing loss compared to the listeners with normal hearing. Despite this, the SEMs remained generally small, indicating high precision of the DIN tests.

**Fig. 9.**
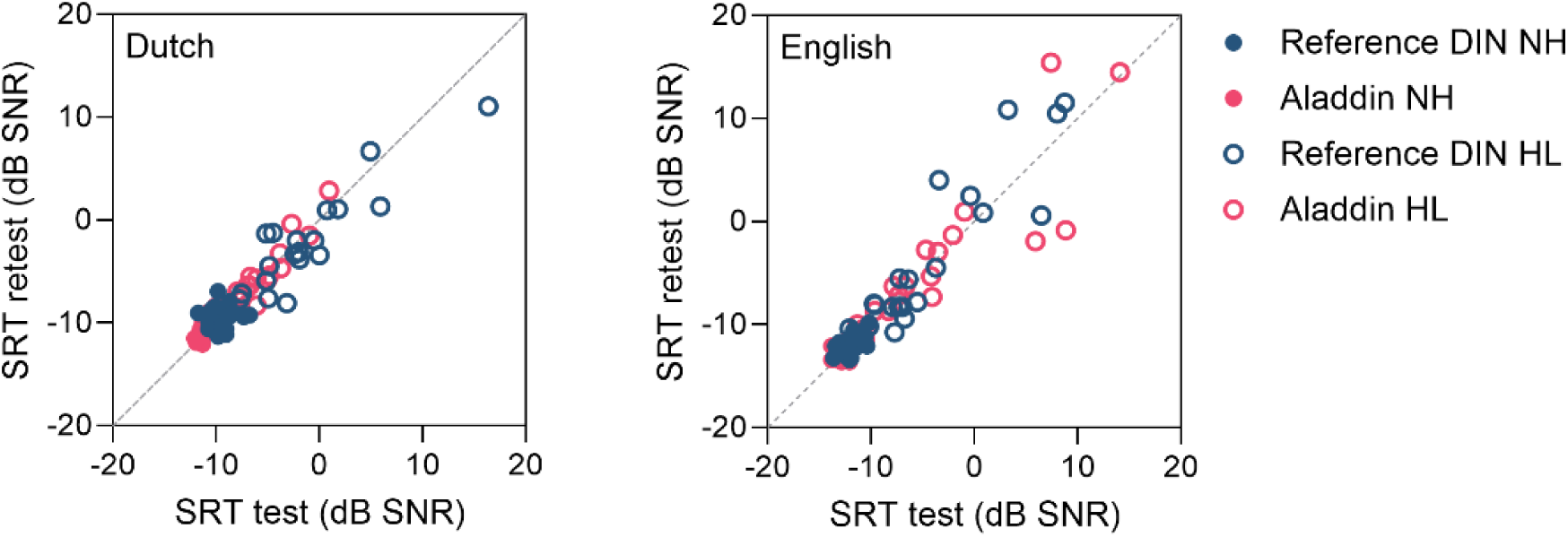
Test-retest SRT scores for the reference DIN test (blue) and the Aladdin test (pink) in Dutch (left panel); and in English (right panel). The filled circles represent the scores of the normal hearing (NH) listeners, the unfilled circles of the listeners with hearing loss (HL).

The screening characteristics of the Dutch and English reference DIN and Aladdin test were evaluated by assessing their ability to discriminate between participants with normal hearing and hearing loss (Figure 10). There was a significant correlation between the SRT and PTA of the better ear for all four tests in the hearing loss group (all p < 0.01). The cutoff SRT was selected as the 95th percentile of the group of listeners with normal hearing. All four tests achieved perfect classification accuracy for all listeners with a hearing loss in the better ear ( >20 dB HL). Note that three listeners in the group with hearing loss had one good ear, so they should not be detected as having hearing loss by the test. The Dutch and English reference DIN tests correctly classified 27 out of 31 listeners with normal hearing. Both Aladdin tests correctly classified 26 out of 31 listeners with normal hearing. This yields specificity and sensitivity of 84% and 100% for both Aladdin tests and 87% and 100% for both reference DIN tests.

**Fig. 10.**
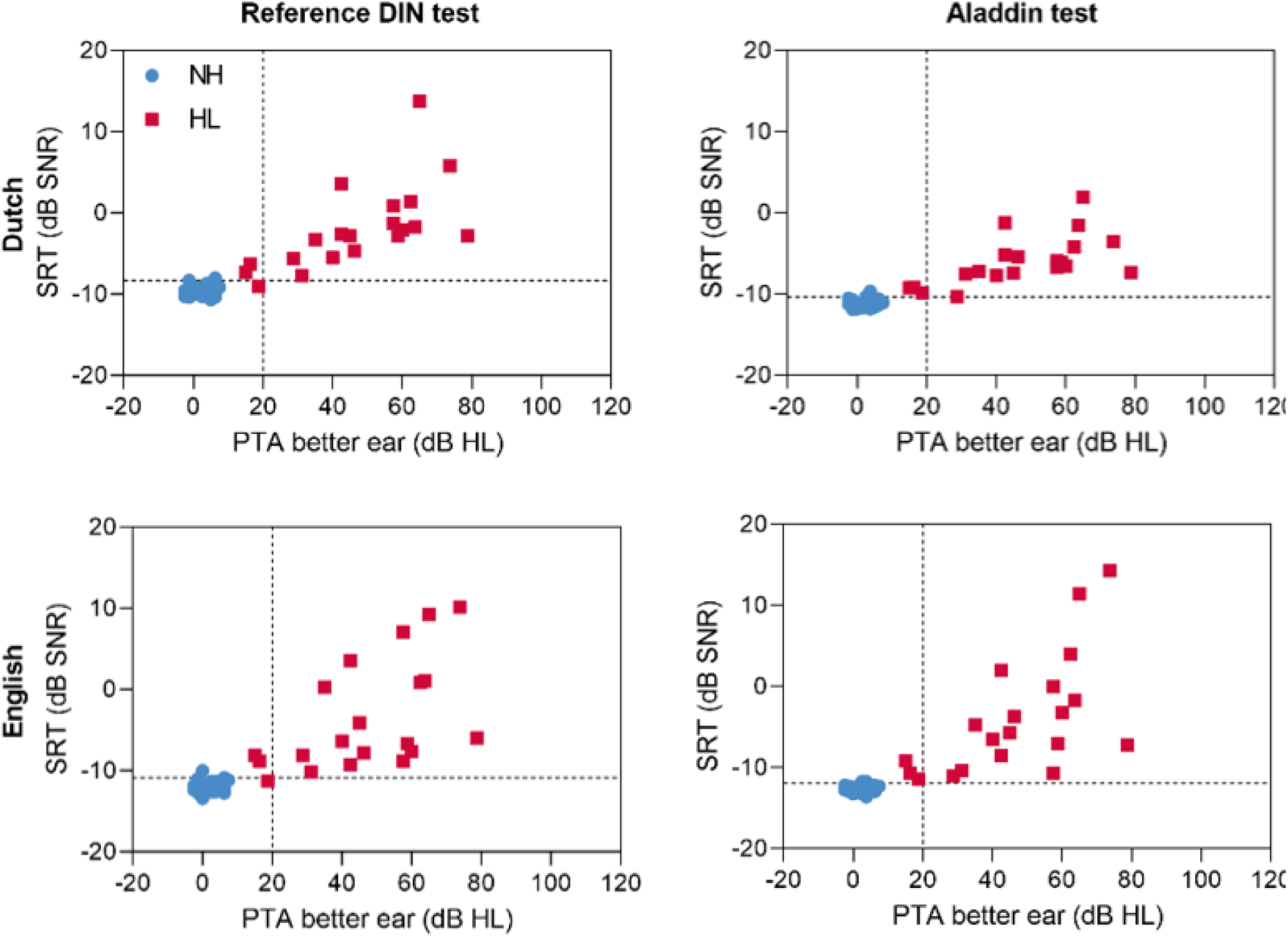
Scatter plots of the SRTs for the reference DIN test (left panel) and the Aladdin test (right panel) in Dutch (top panel) and English (lower panel) as a function of the better ear PTA. Vertical dashed lines at 20 dB HL distinguish between listeners with normal hearing and listeners with hearing loss in the better ear. The horizontal dashed lines denote the SRT values corresponding to the pass/fail criteria for each respective test, determined by the 95th percentile (inclusive) of SRTs within the normal hearing group. The dashed lines divide the plots into quadrants: the lower left and upper right quadrant reflect the correctly classified cases (true negatives and true positives respectively), the lower right quadrant represents false negatives, and the upper left quadrant false positives.

## DISCUSSION

The Aladdin study successfully achieved its objective to generate reliable digits-in-noise (DIN) tests automatically. Utilizing synthetic speech, both Dutch and English versions of the Aladdin test were developed, with digits perceptually equalized through an Automatic Speech Recognition system (ASR). The resulting tests demonstrated similar test characteristics compared to the reference DIN test in their respective languages.

The Aladdin study consisted of four parts. Part I assessed the subjective quality of synthetic speech material in five languages. Results from a listening experiment with native speakers showed good to excellent Mean Opinion Scores (MOS) that did not differ significantly between synthetic and natural speech in any language across various speech materials (digits, words, sentences) and conditions (quiet and noise). These findings align with the results of Part II, where comparisons between natural and synthetic voices were made by assessing speech recognition functions of individual digits presented at fixed Signal-to-Noise Ratios (SNRs) to listeners with normal hearing. Analysis showed no significant differences between natural and synthetic voices in mean DIN Speech Recognition Thresholds (SRTs) or their psychometric slopes, indicating similarity in digit intelligibility across different voices. In contrast, Polspoel, et al. ^23^ found a small but significant difference in mean SRTs and slopes for natural and synthetic words of the same synthetic male and female voices. Note that Part II only considered the evaluation of synthetic Dutch digits. However, it is reasonable to assume that synthetic digits in other languages (e.g., those from Part I) would also yield speech recognition functions comparable to natural digits. This is because Dutch is a relatively less spoken language (approximately 23 million native speakers worldwide), and other more widely spoken languages usually have better coverage by TTS systems (more available voices in Google TTS), potentially resulting in synthetic speech that even more closely resembles human speech.

Findings from both Part I and II indicate that synthetic speech material is suitable for developing hearing tests, consistent with studies that have employed synthetic speech for speech-in-noise tests with digit triplets ^27^, word triplets ^24^, matrix sentences ^28^, and everyday sentences ^25,26^. Polspoel, et al. ^37^ demonstrated that synthetic word lists in quiet are more homogeneously intelligible than commonly used natural lists, without requiring adjustments post-creation of the synthetic words. These results, and the additional benefits of ease of use and affordability, advocate for the use of synthetic speech as the standard approach for creating speech tests.

Presently, Google Cloud TTS supports more than 220 voices across over 40 languages and variants, including various national accents for commonly spoken languages like English (American, British, South-African, Indian, Australian accents). It also includes less commonly spoken languages such as Icelandic, Serbian, Filipino and Hebrew. Although several languages spoken by ethnic groups in low-to middle-income countries are not yet covered by Google TTS, it is anticipated that more languages will be added in the future, as well as regional accents. Moreover, Google TTS already provides a Custom Voice feature, enabling users to train a personalized voice model using their own audio recordings, thereby creating a unique voice. AI advancements even allow for the creation of high-quality synthetic speech across multiple languages utilizing the same voice ^38^. These aspects present a promising avenue for future research aimed at enhancing the Aladdin methodology, thereby facilitating greater comparability of DIN tests across languages and accents.

In Part III, ASRs were evaluated for generating speech recognition functions and determining level corrections for perceptually equalizing digit intelligibility. To establish a human reference for the ASR, level corrections of the natural and synthetic digits were determined based on the listening experiments conducted in Part II. Results showed that the pre-trained open-source ASRs produced SRTs that were significantly higher, and with steeper slopes, compared with human-based evaluations in Part II. The tested systems were not suitable for determining level corrections, likely because their design prioritized minimizing word error rate rather than accurately mimicking human auditory behavior. Consequently, we utilized FADE, an ASR better suited for mimicking human auditory behavior. FADE-derived level corrections demonstrated strong correlations with human-derived level corrections, particularly for synthetic digits. This indicated its effectiveness in the Aladdin approach. Some DIN tests use babble noise ^39,40^, and FADE has been shown to successfully simulate the outcome of speech recognition for different noise maskers, including multitalker babble^41^. Overall, the purpose of FADE is to provide a structured framework for assessing and evaluating auditory discrimination abilities. It was originally designed to predict speech recognition thresholds of the German Matrix sentence test in noise ^34^, and was later extended to predict the outcomes of other speech recognition tests such as DIN and the Göttingen everyday sentences speech test (GÖSA) ^42^. FADE uses no empirical reference and works in different languages and for various noise conditions ^43^. FADE’s predictions in auditory discrimination experiments aligned closely with empirical data for listeners with and without hearing loss ^44,45^, consistent with our findings. To our knowledge, the specific application of determining level corrections for speech material items to achieve perceptual equivalence has not been one of the applications of FADE until now. Note that not all ASRs potentially suitable for the Aladdin procedure were considered in this study. For example, Kaldi, an ASR used for research and development in the field of speech recognition, could possibly also be employed ^46,47^.

In Part IV, the Aladdin test, developed from ASR-based level-corrected synthetic digits, was validated against established reference DIN tests in Dutch and English. Dutch-speaking listeners with normal hearing and with hearing loss performed both reference and Aladdin tests in each language. Strong correlations between Aladdin and reference test SRTs were observed for both Dutch and English versions (r = 0.91 and r = 0.95, respectively). SEM, indicating SRT estimation error, was generally low across all tests. Both Aladdin tests achieved perfect classification accuracy for participants with hearing loss. The SEM and variance in SRTs were noticeably greater among participants with hearing loss compared to those with normal hearing in the English DIN tests (Figure 10). This difference is likely attributable to the older age and lower English proficiency of participants with hearing loss, compared to those with normal hearing. Smits, et al. ^48^ found no significant differences in SRTs between Dutch and English (US) DIN tests among young adult Dutch listeners with normal hearing. Kaandorp, et al. ^49^ demonstrated that linguistic abilities have minimal impact on Dutch DIN test results for young adult university students, suggesting that a basic proficiency in the language is adequate for performing the test. While no significant linear correlation was found between English SRTs and age for either test, participants aged 60 and above displayed significantly greater SRT variability in our study (see supplementary material). Potgieter, et al. ^50^ observed that non-native English speakers with limited self-reported English proficiency performed significantly worse on the South African DIN test compared to both native speakers and non-native speakers with higher self-reported English proficiency. A limitation of our study is the sole assessment of English proficiency through a single yes-or-no question (“Do you have at least a basic understanding of the English language?”), masking variations in proficiency levels and inadvertently including participants with insufficient English proficiency. Ideally, native English participants would have been included in the study.

The Aladdin approach not only simplifies and shortens the test development procedure of a native language DIN test, it also provides a universal guideline for the development of comparable DIN tests in different languages. Currently, comparing DIN test results across languages is not straightforward due to the many variants of the test; including variations in speech spectrum, masking noise, number of trials, antiphasic versus diotic presentation, target audience and test procedures ^4^. With the Aladdin procedure, many of these factors are kept constant. Importantly, the spectra and bandwidth of all Aladdin tests are identical. Thus, “Aladdin” provides a *language-independent* approach to developing DIN tests, making it applicable even in low- and middle-income countries where limited resources and audiometric equipment hinder test development.

In conclusion, the present Aladdin project has demonstrated that valid DIN tests can be generated fully automatically in a uniform manner, without the need for expensive audiometric equipment, professional speakers, and extensive listening experiments. The proposed procedure has a broader application in audiology beyond automating DIN test generation. Conceptually, its methodology could extend to the creation of different speech recognition tests utilizing closed-set stimuli. Examples of such tests are the Matrix sentence test ^51–53^ and Coordinate Response Measure ^54^ but further research is needed to determine whether this approach is indeed suitable for these types of tests.

## METHOD

### Part I: Subjective quality of synthetic (text-to-speech) speech material for hearing tests

#### Material

After assessing multiple text-to-speech (TTS) systems for generating speech material, Google Cloud TTS emerged as the preferred choice. Its selection was based on its superior speech quality, well-documented technology, extensive range of voices and languages, and an API that streamlines bulk speech production. Currently, Google Cloud TTS supports over 40 languages and offers more than 220 voices by leveraging powerful neural networks known as WaveNet through its API ^29^. Dutch voices were chosen based on naturalness by two speech therapists from the Amsterdam UMC, while voices for other languages were selected by the first author based on perceived speech quality. Google Cloud TTS produced 16-bit WAV files for digits, words, and sentences at a 44.1 kHz sample rate. Our consideration of multiple languages aligns with Aladdin’s goal to automate hearing test creation for any language.

The speech material, created in five languages (Dutch, English, French, Spanish, and Mandarin), included exact replications of digits, words, and sentences used for standard speech audiometry. Examples of standard speech materials in the Netherlands are the original DIN test digits^6^, NVA words^55^, and VU-98 sentences^56^. The corresponding countries where the speech material is used are the Netherlands, USA, France, Spain, and China. Supplementary material provides an overview of the speech material types used in each country and the synthetic Google voices used for their production. All material was created in both a male and female synthetic voice. Additionally, the experiment included both natural and degraded voices, wherein the natural voice represented the standard speech material and served as a benchmark for the synthetic voices. The degraded voice was a 12-channel noise-carried vocoded voice to cover the entire range of the scale.

A listening experiment, following International Telecommunication Union - Telecommunications sector (ITU-T) P-series recommendations for speech quality assessment^20^, was conducted to evaluate subjective quality across various speech materials and languages. Self-developed software facilitated listeners, with different mother tongues, to select their native language, rate audio sound files in quiet and noise, and answer the question: “Overall impression - how would you rate the quality of the speech you just heard?” (with “(ignore the noise)” added for sound files in background noise). Ratings were provided on a 5-point Mean Opinion Score (MOS) scale, ranging from “Excellent” (5 points) to “Very poor” (1 point) ^30^. Official translations of the scale were utilized, and translations of the “Overall impression” question were authenticated by native speakers. All translations are available in the supplementary materials. The software instructions were presented in English.

The software was installed on a laptop (Dell Latitude 5580 Core 15-6300U, 16 GB) provided with headphones (Sennheiser HD 280 Pro). Participants were asked to complete a 15-minute speech rating task on it. The laptop was calibrated so that all speech sound files were presented diotically at a level of 65 dB SPL.

#### Participants

Fifty native speakers participated: 10 Dutch (mean age 38 ± 12 years), 10 American (M = 37 ± 8 years), 10 French (M = 27 ± 1 years), 10 Spanish (M = 36 ± 5 years) and 10 Chinese (Mandarin; M = 31 ± 7 years). The Dutch participants were recruited by the first author of this paper. Cochlear, a partner in this study, assigned an American, French, Spanish, and Chinese contact person to recruit and instruct 10 participants (5 male, 5 female) each in the USA, France, Spain, and China, respectively. Contacts were instructed to select individuals under 50 years old with no self-reported hearing problems and basic proficiency in English to understand the software instructions. Most participants were Cochlear employees, some were acquaintances of the contact people. A calibrated laptop was sent to Cochlear offices in the different countries. After obtaining test results for 10 participants, the equipment was returned to the first author and then sent to the next contact person in a different country. This process occurred from October 2021 to July 2023. The VUmc medical research ethics committee determined that the study did not require additional ethics evaluation (protocol number: 2021.0060).

#### Testing procedure

The design of the speech assessment task involved within-subject repeated measures. Participants, guided by a contact person, completed the test independently on a laptop in a quiet environment. Using the software, participants selected language, age, gender, and indicated normal hearing. The test comprised 6 sections, with participants rating sound files on a 5-point MOS scale^30^. Sections were presented in a fixed sequence and included: (1) digits in quiet, (2) digits in noise (3) words in quiet (4) words in noise (5) sentences in quiet and (6) sentences in noise. Each section featured four voices (natural, synthetic male, synthetic female, degraded), each presented randomly four times. Consequently, there were 16 presentations to evaluate in each section, comprising four practice runs with the four voices at the onset of each segment to familiarize the participant with the task (these were not scored). Each voice in each section was scored three times per participant, and these scores were averaged. Sections (1) and (2) comprised three digit triplets per presentation, (3) and (4) had six words, and (5) and (6) featured three sentences per presentation, resulting in each presentation lasting around 5 to 6 seconds. Participants could take breaks between sections. Each voice type was presented three times in each section. Average scores per section per voice were calculated after collecting all speech rating data, adhering to standard MOS regulations ^30^. The experiment had a total duration of approximately 15 minutes.

#### Statistical analysis

The Kruskal-Wallis test compared scores of synthetic and natural voices across languages for digits, words, and sentences. It serves as a non-parametric alternative to the F-test due to violations of the normality assumption of the dependent variable.

### Part II: Comparing the speech recognition functions of natural and synthetic digits

#### Materials

The synthetic (female and male voice) and natural Dutch digits (0 to 9) from Part I were used for further evaluation. The natural digits and masking noise were adopted from the standard Dutch DIN test before any level corrections were applied^6^. These natural digits were pronounced by a male professional speaker. While recommending the use of female voices for future automatically generated DIN tests (’Aladdin-tests’) for consistency, this study also included a male synthetic voice for better comparison with the natural male voice.

The average spectrum of the synthetic digits (long-term average speech spectrum, LTASS) was adjusted to match the ‘idealized speech spectrum’ from the speech intelligibility index (SII) standard: a constant sound pressure spectrum level from 100-500 Hz and decreasing from 500-9500 Hz at a rate of 9 dB per octave. The LTASS adjustment involved correcting the synthetic speech spectrum by the difference between the LTASS of the synthetic and the standard LTASS using PRAAT ^57^. This ensures a uniform speech (and noise) spectrum for all future Aladdin test materials, preventing differences in LTASS from influencing intelligibility variations between Aladdin tests. Note that the masking noise for all Aladdin tests is identical, with its spectrum mirroring the idealized LTASS. The spectra of the natural digits and the corresponding noise were left unadjusted. The LTASS of the male synthetic digits and noise was aligned with natural DIN material using PRAAT in a similar manner. Subsequently, the root mean square (RMS) levels of all synthetic and natural digits were equalized. Following the approach outlined by Smits, Theo Goverts and Festen ^6^, 120 unique digit triplets were constructed derived from the 10 digits in each voice, with each triplet containing three distinct digits. Silent intervals of 500 ms preceded the first digit and followed the final digit, while 150 ms intervals separated the digits.

The testing equipment for the listening task included Sennheiser HDA200 headphones connected to a Dell Optiplex780 PC via a Sound Blaster Creative soundcard (THX).

#### Estimating the Speech recognition function

##### Participants

Twenty-four native Dutch-speaking adults (M = 21 ± 2 years) with normal hearing participated. All participants had pure-tone thresholds ≤20 dB HL across octave frequencies from 0.25 to 8 kHz in the test ear. Only one ear was tested in the subsequent speech recognition experiment. The mean pure-tone average (PTA) at 0.5, 1, 2, and 4 kHz of the tested ears was 3.1 ± 2.9 dB HL. Left and right ears were alternated between participants, unless only one ear qualified. Recruitment occurred on the university campus, with participants receiving compensation and providing written informed consent. The VUmc medical research ethics committee declared that the study did not need further ethics evaluation (protocol number: 2020.0758).

##### Testing Procedure

The experimental study employed a within-subject repeated measures design. Testing took place in a sound-treated booth and stimuli were monaurally presented to the listener via headphones. DIN tests were presented at fixed SNRs of -14, -12, -10, -8, -6, and 0 dB, with an overall presentation level set at 65 dB SPL. Participants were divided equally, with one-third starting with the female synthetic DIN test, one-third with the male synthetic DIN test, and one-third with the male natural DIN test. Each test comprised 120 triplets organized into 6 lists of 20 triplets, ensuring each digit appeared 6 times per list. Participants encountered one list per presentation level for each DIN test, maintaining consistent presentation numbers per digit and signal-to-noise ratio (SNR) for speech recognition function estimation. The order of the 6 lists remained fixed for each participant across the three DIN tests to minimize learning effects. To mitigate fatigue, the SNR order alternated between easier (-4, -6, and 0 dB SNR) and more challenging (-14, -12, and -10 dB SPL) conditions. Presentation levels remained fixed for all three DIN tests within each participant. Each DIN test began with a practice list to minimize potential learning effects.

The participants were verbally instructed to repeat three digits after each trial and encouraged to guess if uncertain. When they could not understand one or more digits, they could say “blank” (e.g., “4, blank, 7”). In data analysis, blanks were replaced with a random digit, resulting in a 10% guess rate per digit. Stimuli were not repeated, and no feedback was given.

### Part III: Determining level corrections from ASRs

Speech recognition functions of the Dutch and English natural and synthetic digits from Part I were estimated via ASRs. Various commercial and open-source ASRs were considered for the Aladdin procedure, including three pre-trained cloud-based ASRs and one non-pre-trained ASR (FADE). The resulting speech recognition functions were compared to those obtained by humans in Part II, and ASR-based level corrections were determined.

#### Materials & Methods

##### Cloud-based ASRs

Three cloud-based ASRs were assessed: Google Cloud ASR ^31^, Microsoft Azure Bing Speech API ^32^, and IBM Watson Speech to Text ^33^. These systems are typically trained on extensive datasets, with Google Cloud’s ASR, for instance, trained on millions of hours of audio and billions of text sentences, supporting over 110 languages. Utilizing advanced deep learning neural network algorithms, these systems convert speech to text and offer real-time processing and customization through APIs. Despite their similarities, these systems vary in target applications, data processing, model training, technology, training data, and supported languages.

The spectrum-adjusted natural male and synthetic female Dutch digits, along with corresponding noise from Part II, formed the basis for creating 120 distinct digit triplets in noise across 24 SNRs (-4.5 to 7 dB SNR in 0.5 dB steps). The digit triplets in noise were evaluated across all three ASRs via an API in Python, yielding scores ranging from 0 to 100% for most digits. The output was standardized to always consist of 3 characters, replacing non-digit responses with blanks and converting words to digits where applicable. Single-digit responses were correctly positioned, with the remaining characters as blanks, later substituted with random digits (0-9) to introduce a 10% guessing chance per digit presentation. Additional triplets with lower SNRs (-5 to 7.5 dB SNR in 0.5 dB steps) were created for the natural voice, specifically for Google Cloud and Azure ASR, due to overly high scores and lack of speech recognition function estimations. For the analysis of speech recognition functions per digit, only SNRs yielding scores between 20 and 90% were considered.

##### FADE ASR

FADE is a Hidden Markov Model based model that simulates the human speech recognition process ^19,34,35,43,45^, and has demonstrated in previous research to accurately predict SRTs of various speech materials ^42,58^. It stands for “simulation Framework for Auditory Discrimination Experiments” and uses front-end auditory spectro-temporal features for the initial processing of the audio signals before further analysis. Options include methods such as the ‘Gabor filter bank (GBFB)’ and ‘Mel-Frequency Cepstral Coefficients (MFCC)’ to resemble human auditory processing. FADE operates with a distinct training and testing phase. During the training phase, the system, utilizing hidden Markov models (HMMs), is exposed to labeled audio data to learn patterns and features associated with different stimuli. Techniques like feature extraction are used to capture relevant characteristics. Parameters are adjusted based on feedback from the training data to improve performance. In the testing phase, the trained system is evaluated on unseen data to assess its ability to discriminate between stimuli. This iterative process allows researchers to develop and refine algorithms for tasks such as speech recognition or sound classification, enhancing the system’s ability to generalize to new data.

The FADE source code and instructions are available on GitHub^59^ and were installed on a Linux OS (Ubuntu). We included the spectrum-adjusted Dutch natural male and synthetic female digits from Part II, along with spectrum-adjusted synthetic English female digits. The LTASS of the English digits mirrored that of the Dutch synthetic voice, with an LTASS that matches the standard speech spectrum (see Part II). The three sets of digits were considered separately. During this phase, English synthetic digits were selected alongside Dutch digits, as the subsequent development of the Aladdin test also included an English version. Additionally, Dutch listeners generally have sufficient familiarity with English to complete a DIN test, unlike with other languages such as French, Spanish, or Mandarin.

FADE was trained on each set of digits in speech-shaped noise at various SNRs (-24 to 6 dB SNR, default), with steps of 1 dB. To enhance precision in estimating the speech recognition threshold, we adjusted the default step size from 3 dB to 1 dB. We set both training and test samples to 1000 and compared both GBFB and MFCC front-end feature. Because the LC determined from using GBFB showed stronger correlation with human results, we decided to use GBFB as the standard. This decision aligns with Schädler, Meyer and Kollmeier ^60^, who preferred GBFB over MFCC due to its enhanced robustness against additive noise. Following the initial FADE run, the output yielded binary results (0 or 1), indicating whether a digit was correctly recognized by the ASR or not, at every SNR for an optimal training SNR. A training SNR of -9 dB SNR was selected as optimal after considering different possibilities. This facilitated the estimation of speech recognition functions for each digit. Then, the level-corrected digits were run through FADE again to generate new speech recognition functions. When all digits reached a score of 80% at 6 dB SNR, the level corrections were determined by subtracting the individual SRT of that digit from the average SRT. Then, the overall speech recognition function was determined, and the SRT and slope were noted. Next, the corrected digits were run through FADE again, and new level corrections were determined along with the overall speech recognition function. This process continued until the slope of the overall speech recognition function did not improve by more than 1%-point/dB. The accumulated level corrections from all runs formed the final corrections of the FADE system, which were then compared to the human-based corrections in Part II. The Root mean square (RMS) error of the level corrections was calculated by taking the square root of the average of the squared differences between the human-based and FADE ASR-based level corrections.

### Part IV: Comparing the Aladdin test to the original DIN test

#### Material

Participants in the study performed four types of DIN tests: a Dutch reference DIN test, an English reference DIN test, a Dutch Aladdin test, and an English Aladdin test. The reference Dutch DIN test was based on Smits, Theo Goverts and Festen ^6^ ‘s original DIN test, featuring natural male digits that had been level-corrected through a human listening experiment. The reference English DIN test was the one by Motlagh Zadeh, et al. ^61^, employing level-corrected natural female digits spoken by a Midwestern American English speaker. Both the Dutch and English Aladdin tests were developed using spectrum-adjusted synthetic digits that had been level-corrected by the FADE ASR. The LTASS of both sets of Aladdin digits, along with the spectrum of the corresponding noise, matched the standard LTASS. Furthermore, the assembly of these tests followed the methodology established by Smits, Theo Goverts and Festen ^6^, as detailed in Part II.

#### Participants

Twenty-eight listeners with normal hearing (NH, M = 25.5 ± 4.5 years) and 20 listeners with hearing loss (HL, M = 59.2 ± 14.2 years) participated in the listening experiment. Pure-tone audiometry was conducted using a standard clinical audiometer (Decos audiology) and Sennheiser HDA200 headphones. Listeners with normal hearing had pure-tone thresholds of ≤20 dB HL across all octave frequencies from 250 Hz to 8 kHz in both ears and were recruited from the University campus. Listeners with hearing loss were recruited through flyers at the Amsterdam UMC hospital audiology department and a post on a Facebook group for hearing-impaired individuals. All participants were native Dutch speakers with at least basic knowledge of English, received compensation, and provided written informed consent. The VUmc medical research ethics committee deemed further ethics evaluation unnecessary (protocol number: 2023.0202).

#### Testing Procedure

The study used a within-subject repeated measures design. Participants completed the DIN tests independently, seated in a sound-treated booth with a computer, without a test administrator present. Stimuli were presented diotically through Sennheiser HDA200 headphones connected to a Dell Optiplex780 PC via a Sound Blaster Creative soundcard (THX). The order of the four types of DIN tests was counterbalanced using a Latin square design across all participants. Each test involved presenting 24 digit triplets in a one-down, one-up adaptive manner, similar to the original DIN presentation method ^48^. The SRT was determined as the average SNR of the last 20 out of 24 presentations. They performed each type of test twice to assess test-retest accuracy. Additionally, the first condition was repeated once as a practice session to familiarize participants with the task. The overall presentation level was fixed at 65 dB HL for the listeners with normal hearing. The listeners with hearing loss had their presentation level adjusted individually for comfort, with an examiner presenting digits in quiet and adjusting the level until comfort was reached prior to the experiment. Thirteen out of twenty listeners with hearing loss were presented with levels exceeding 65 dB SPL (with an average of 87.5 dB SPL).

## ACKNOWLEDGEMENTS

We acknowledge the contributions of the following individuals to our research: Hans van Beek for software development; Filiep Vanpoucke for facilitating outreach via Cochlear, and all international contacts for participant recruitment in our multilingual experiment; and Marc Schädler for the assistance in setting up FADE. David Moore is supported in part by the NIHR Manchester Biomedical Research Centre (UK). We acknowledge the hearWHO language development partners for using the original DIN recordings in Part I. This study was supported by Health∼Holland, Top Sector Life Sciences & Health

## AUTHOR CONTRIBUTIONS

C.S. acquired funding, conceived the project, and designed the experiments. S.P. performed the experiments, analyzed the data and wrote the manuscript with input from all authors. D. R. M., D. S. and S.E.K. substantially revised the manuscript. All authors have approved the submitted version.

## DATA AVAILABILITY

The data that support the findings of this study are available from the corresponding author upon reasonable request

