## supplementary material for "Global access to speech hearing tests"

**Supplementary material (1):** Overview of the speech material types used in each country and the male (M) and female (F) synthetic Google voices used for their production.

| | Dutch | English | French | Spanish | Mandarin |
|---|---|---|---|---|---|
| Digits | Digits of the DIN test (Smits, Theo Goverts, & Festen, 2013) | hearWHO digits (Potgieter et al., 2018) | hearWHO digits (De Sousa et al., 2022) | hearWHO digits (De Sousa et al., 2022) | hearWHO digits (De Sousa et al., 2022) |
| Words | NVA word lists (Bosmana & Smoorenburg, 1995) | NU-6 (Tillman & Carhart, 1966) | Fournier disyllabic words (Fournier, 1951) | Test de Navarram developed by Clinica Universitaria de Navarra (personal communication) | Mandarin PB monosyllable speech test (Fei et al., 2010) |
| Sentences | VU-98 sentences (Versfeld et al., 2000) | American English Hearing in Noise Test (Vermiglio, 2008) | MBAA2 (James et al., 2023) | Sharvard Corpus (Aubanel, Lecumberri, & Cooke, 2014) | HOPE Mandarin sentences (Xi et al., 2012) |
| Synthetic Voices | M**:** nl-NL Wavenet B<br>F: nl-NL Wavenet D | M: en-US Wavenet B<br>F: en-US Wavenet F | M: fr-FR Wavenet C<br>F: fr-FR Wavenet D | M: es-ES Wavenet B<br>F: es-ES Wavenet C | M: cmn-CN Wavenet B<br>F: cmn-CN Wavenet A |

**Supplementary material (2):** Translations of the "Overall impression" question and MOS scale of the speech rating experiment in Part I.

**English**

Overall impression – How do you rate the quality of the speech of what you just heard? (ignore the noise)

- Excellent
- Good
- Fair
- Poor
- Very Poor

**Dutch**

Algehele indruk - Hoe beoordeelt u de kwaliteit van de spraak van wat u zojuist hebt gehoord? (negeer de ruis)

- Uitstekend
- Goed
- Redelijk
- Matig
- Slecht

**French**

Impression générale - Comment évalueriez-vous la qualité de la parole de ce que vous venez d'entendre? (ignorez le bruit)

- Excellente
- Bonne
- Passable
- Médiocre
- Mauvaise

**Spanish**

Impresión general - ¿Cómo calificaría la calidad del discurso que acaba de escuchar? (ignore el ruido).

- Excelente
- Buena
- Regular
- Mediocre
- Mala

**Mandarin**

总体印象 - **您如何**评价刚刚听到的语音质量？（**忽略背景噪音**）

- 很好
- 好
- 一般
- 差
- 很差

**Supplementary material (3):** Level corrections (LC) of the natural male and synthetic female Dutch digits derived from the listening experiment with human listeners from Part I, as a function of the LC derived from Google ASR (left panel), the Microsoft Azure ASR (middle panel) and the IBM Watson ASR (right panel) (Part III). Three significant correlations ($p$ < 0.05) were found, indicated by *.

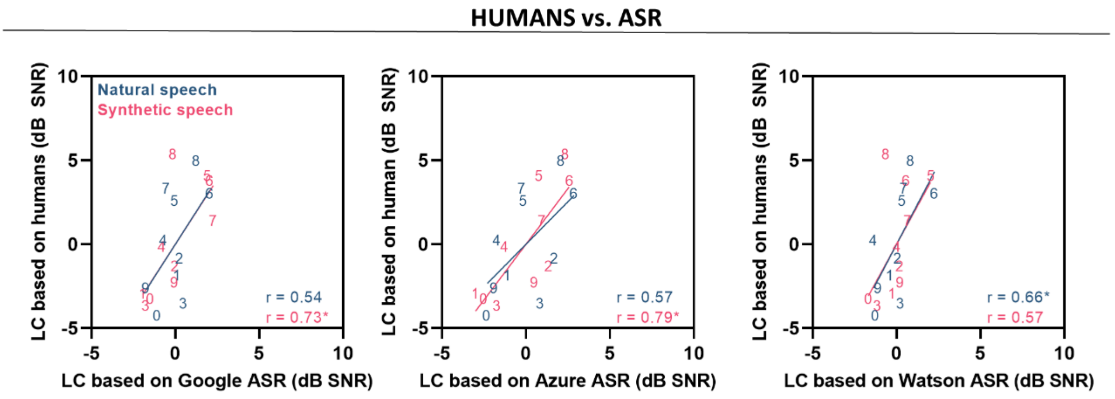


**Supplementary material (4):** Scatterplots of SRTs as a function of age for the Dutch and English DIN and Aladdin test (Part IV). Participants aged 60 and above displayed significantly greater SRT variability than listeners under 60 for the English reference DIN (independent t-test, equal variances not assumed, t(15.168) = -2.082 , $p = 0.05$)

**SRT vs. AGE**

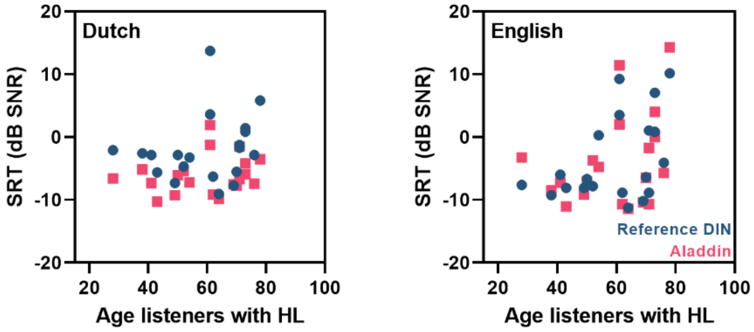
